# White Matter Integrity Correlates with Strength of Response to Deep Brain Stimulation in Treatment-Resistant Obsessive-Compulsive Disorder

**DOI:** 10.64898/2026.05.11.26352565

**Authors:** Grace Leslie T. Nitcheu, Reem El Jammal, Hideo Suzuki, Sarah Soubra, Thomas A. Hamre, Melissa A. Ryan, Saipravallika Chamarthi, Vinayak Belavadi, Zachary Perry, Thomas P. Kutcher, Victoria Gates, Garrett P. Banks, Nora Vanegas Arroyave, Eric A. Storch, Wayne K. Goodman, Sameer A. Sheth, Sarah R. Heilbronner, Nicole R. Provenza

## Abstract

**Background:** Deep brain stimulation (DBS) is effective for approximately two-thirds of patients with treatment-resistant obsessive-compulsive disorder (OCD). While prior work has emphasized the engagement of specific white matter tracts in mediating outcomes, the contribution of region-specific white matter integrity to clinical response remains unclear.

**Methods:** Twelve patients with treatment-resistant OCD underwent preoperative neuroimaging and DBS at our center. We assessed OCD severity preoperatively and at ∼18 months postoperatively. We extracted mean fractional anisotropy (FA) for the anterior limb of the internal capsule (ALIC) and a control tract and used Spearman’s rank correlations to evaluate associations between FA and symptom improvement. We additionally evaluated this relationship for 49 white matter bundles. Finally, we used diffusion tractography to determine endpoints connected with ALIC voxels most predictive of symptom improvement.

**Results:** Higher preoperative ALIC FA was associated with greater clinical improvement following DBS (p=0.002). This effect was specific to the ALIC and not the control tract. Hemispheric asymmetry (right>left) in ALIC FA was moderately correlated with clinical improvement. Among all 49 bundles, the right ALIC demonstrated the strongest association with clinical improvement. Streamlines passing through the ALIC voxels that most strongly correlated with outcome ended in the diencephalon and superior frontal cortex.

**Conclusions:** Baseline structural integrity of the ALIC was associated with the magnitude of clinical improvement following DBS for OCD. These findings suggest that regional variation in white matter integrity may reflect an underlying circuit disruption amenable to DBS, supporting the use of neuroimaging-based metrics as potential biomarkers in DBS treatment.

## Background

Obsessive-compulsive disorder (OCD) is a debilitating psychiatric disorder with a lifetime prevalence of 1–3%, characterized by intrusive, recurrent thoughts (obsessions) and repetitive behaviors or mental rituals (compulsions) performed to reduce distress. Conventional treatments, including selective serotonin reuptake inhibitors and cognitive-behavioral therapies such as exposure and response prevention, are effective for approximately 50–70% of patients; however, an estimated 25-40% remain refractory to these interventions [1,2]. In this treatment-refractory population, deep brain stimulation (DBS) of the ventral capsule/ventral striatum (VC/VS) is effective in ∼66% of patients [1,3].Despite overall favorable outcomes, a subset of patients do not meet response criteria (≥35% reduction in the Yale-Brown Obsessive Compulsive Severity Scale [Y-BOCS]) even after prolonged therapy, highlighting substantial heterogeneity in treatment response.

In recent years, efforts to explain this variability have focused on identifying the specific white matter tracts acting as mediators of clinical outcome, informing refinements in lead placement and targeting strategies [4,5]. However, the contribution of regional white matter integrity itself remains poorly understood. Diffusion-weighted magnetic resonance imaging (dMRI) provides several scalar metrics of white matter structure, including fractional anisotropy (FA), which indexes white matter integrity. In this study, we used dMRI to examine regional and hemispheric associations between preoperative FA and clinical response to DBS for OCD.

## Methods

We retrospectively analyzed preoperative dMRI data from 12 patients with treatment-refractory OCD who underwent DBS targeting the VC/VS at our institution. We assessed clinical outcomes using the Yale-Brown Obsessive-Compulsive Scale (Y-BOCS) at baseline and at the closest available follow-up to 18 months following stimulation activation.

We acquired preoperative diffusion-weighted images (DWIs), using single-shell high-angular-resolution for six subjects (voxel size=2mm isotropic; 19 b0 images with AP and PA phase encoding; 256 directions atb=1000s/mm^2^ with AP phase encoding; TR/TE=4230/65ms) or multi-shell, multi-band for the other six subjects (voxel size=1.5mm isotropic; 14 b0 images; 46 directions per shell at b=1250 and 2500s/mm^2^; AP and PA phase encoding; TR/TE=3200/87ms), on a 3T Siemens MRI scanner using a 32-channel head coil. Then, we preprocessed DWIs for distortion corrections and created FA maps using MRtrix3. Furthermore, these FA maps were skeletonized using tract-based spatial statistics (TBSS) from FSL.

We first performed region-of-interest (ROI) analyses in native diffusion space. We registered the JHU ICBM-DTI-81 white matter atlas from MNI space to each subject’s FA map using rigid-body registration in FSL FLIRT (6 degrees of freedom, correlation-ratio cost function). We then applied the resulting transform to the JHU label atlas using nearest-neighbor interpolation. Using these subject-specific atlas labels, we extracted mean FA from the anterior limb of the internal capsule (ALIC), the main white matter bundle at the VC/VS site. We also extracted FA from the posterior thalamic radiation (PTR), which is anatomically distant from both the DBS lead and the ALIC, as a negative control. We evaluated associations between ROI FA and clinical outcome using Spearman’s rank correlations with corresponding 95% confidence intervals (CIs). We additionally quantified hemispheric FA asymmetry as (right − left)/(right + left).

We next evaluated whether ALIC FA demonstrated the strongest association with outcome relative to other white matter bundles. In this case, we were concerned about partial voluming effects, so we masked each white matter bundle in the JHU atlas by the white matter skeleton to extract mean FA for the skeleton of each tract separately in the left and right hemispheres. Using Spearman’s rank correlation, we then assessed strength of associations between tract-specific FA and percent Y-BOCS reduction. (These were not evaluated for significance due to concerns about circular analysis [4].) In follow-up analyses, we used partial Spearman’s rank correlation while controlling for patient age or global skeletonized FA.

Finally, to identify foci within the ALIC of which white matter integrity was most strongly associated with outcome, we evaluated voxel wise associations between FA and percent Y-BOCS reduction within the ALIC mask using nonparametric permutation testing with threshold-free cluster enhancement. We used a thresholded map of high-effect voxels (97%) to define a region-of-interest (ROI) in each hemisphere for tractography. We then performed deterministic tractography in DSI Studio using quantitative anisotropy–based tracking, seeding from this ROI and utilizing a normative connectome in DSI Studio ([5]). We quantified cortical and subcortical endpoints by intersecting streamline endpoints with the Desikan-Killiany-Tourville (DKT) atlas in MNI space.

## Results

Structural integrity of the ALIC (computed via FA) showed a strong and significant positive correlation with clinical improvement (% Y-BOCS reduction) (Left: ρ=0.685, 95% CI [0.164, 0.914], p=0.0173; Right: ρ=0.832, 95% CI [0.451, 0.949], p=0.0014 (Figure 1A). In contrast, FA of the control tract, the PTR, was not significantly associated with clinical outcome (Left: ρ = 0.385, 95% CI [-0.328, 0.871], p=0.218; Right: ρ=0.224, 95% CI [-0.409, 0.736], p=0.485). Hemispheric asymmetry in ALIC FA was also significantly correlated with clinical improvement (ρ=0.629, 95% CI [0.039, 0.907], p=0. 0324), indicating greater rightward FA asymmetry in patients with stronger treatment response (Figure 1A). Unlike change in Y-BOCS score with treatment, baseline Y-BOCS score was not significantly related to ALIC FA (Left: ρ= -0.219, CI [-0.888, 0.565], p= 0.4931; Right: ρ=-0.421, 95% CI [-0.927, 0.248], p=0.1726). Additionally, associations between ALIC FA and percent Y-BOCS reduction remained significant after controlling for DWI acquisition protocol (ρ=0.821, 95% CI [0.361, 0.968], p=0.0011), indicating that findings were not driven by differences in imaging parameters.

**Figure 1.**
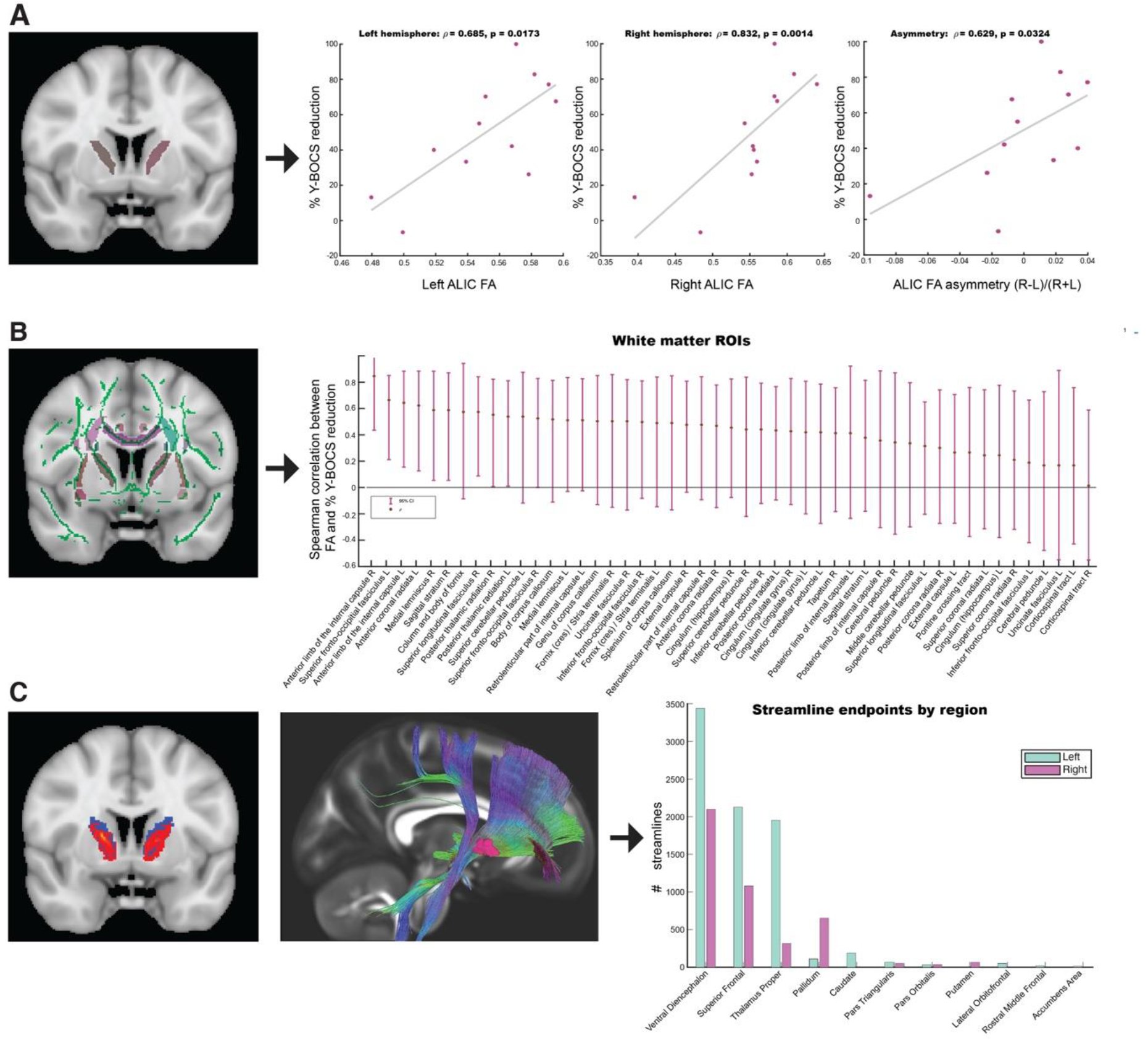
White matter integrity correlates of clinical response to DBS for OCD. **A**. Correlations between baseline ALIC FA and percent Y-BOCS reduction analyzed separately for the left and right hemispheres, as well as hemispheric asymmetry (R-L/(R+L). **B**. When evaluating how 49 skeletonized bundles predict percent Y-BOCS reduction, the right ALIC demonstrated the strongest association with clinical outcome. On the left, green indicates skeletonized white matter, and other colors show bundles; the intersection was used for this analysis to reduce partial voluming effects. (**C**) Distribution of normative tractography endpoints seeded from ALIC subregions most strongly associated with outcome. Streamline endpoint voxel counts are shown for cortical and subcortical regions defined by the DKT atlases, plotted separately for the left and right hemispheres. Abbreviations: ALIC, anterior limb of the internal capsule; FA, fractional anisotropy; L and R denote left and right hemispheres.

In an atlas-based analysis of skeletonized FA using the JHU ICBM-DTI-81 white matter atlas, baseline FA of the right ALIC demonstrated the strongest association with percent Y-BOCS reduction (Figure 1B). This was also true when controlling for global FA or age (Supplement).

To characterize the putative anatomical connectivity of ALIC subregions most strongly associated with outcome, tractography was performed using voxels exhibiting the highest FA– outcome correlations. Endpoint analyses revealed prominent projections to the superior frontal cortex as well as subcortical connections involving the thalamus and ventral diencephalon, and weaker projections to the basal ganglia and ventral prefrontal cortex (Figure 1C).

## Conclusions

In this study, preoperative white matter integrity of the ALIC was associated with the magnitude of clinical improvement following DBS for treatment-refractory OCD. Across analytic approaches, FA of the right ALIC in particular demonstrated a strong, consistent relationship with percent reduction in Y-BOCS scores. The ALIC is also the target white matter bundle for VC/VS DBS, suggesting that baseline microstructural properties of to-be stimulated pathways may help determine DBS efficacy or candidacy.

One question unanswered by our work is why right ALIC FA is more strongly associated with treatment response than left; moreover, hemispheric asymmetry itself is predictive of outcome. Microscopy studies with postmortem tissue have demonstrated organizational differences in the right vs left ALIC. Specifically, frontopontine bundles on the left are smaller and more numerous than those on the right [6]; how such results link to commonly observed hemispheric asymmetries in DBS for OCD (e.g., [7]) is unclear.

Prior work across movement and psychiatric disorders has demonstrated that preoperative neuroimaging features can predict clinical response to neuromodulatory and surgical interventions. In Parkinson’s disease, resting-state functional connectivity between the globus pallidus internus (GPi) and subthalamic nucleus (STN), the two established DBS targets for motor symptoms, has been shown to predict motor response [8]. These findings highlight the importance of baseline circuit organization and individual anatomical variation in determining treatment response.

Diffusion metrics have also been used to predict motor recovery after transcranial direct current stimulation during physical therapy [9]. Higher FA in the corticospinal tract was correlated with greater motor recovery. This suggests that greater connectivity between the cortex and the rest of the motor pathway enabled stimulation to produce greater improvements. As VC/VS DBS primarily targets white matter fibers traversing the ALIC, it follows that patients with more robust, highly myelinated connectivity between the frontal lobes and subcortical structures could similarly achieve greater improvement from stimulation. In patients with greater ALIC FA, DBS may have a greater capacity to positively affect frontal lobe function.

Neuroimaging biomarkers have also been used to anticipate response to neuromodulation treatment in OCD [10]. For DBS, metabolic activity in the subgenual anterior cingulate cortex correlates with eventual response [11]. Aside from DBS, preoperative metabolism within the posterior cingulate cortex predicts degree of Y-BOCS improvement following cingulotomy [12]. A right anterior cingulate ROI is significantly associated with cingulotomy outcome (lower gray matter volume associated with improved response) [13]. Greater preoperative dorsal anterior cingulate connectivity to the dorsal caudate is also associated with greater response to capsulotomy [14]. Together, these studies not only support the broader concept that preoperative circuit characteristics influence surgical outcomes in OCD, but implicate many of the same circuits identified here, as the anterior cingulate cortex is a strong contributor of fibers to the ALIC. Along with conflicting reports of abnormal FA in the ALIC of OCD patients [15,16], our findings emphasize interindividual variability within a DBS-treated cohort and suggest that higher preoperative FA, particularly within the right ALIC, may function as a predictor of therapeutic responsiveness.

## Data Availability

All data produced in the present study are available upon reasonable request to the authors

## Contributorship

GTN was involved in the study design, performed the statistical analyses, interpreted the data, and wrote the manuscript. REJ, NVA, MAR, and HS were involved in data analysis and provided critical revisions of the manuscript. SS, TAH, and SC were involved in data processing and provided critical revisions of the manuscript. TPK, VG, VB, and ZP were involved in data collection and processing. GPB, EAS, and WKG were involved in data collection, supervision, and contributed critical revisions of the manuscript. SRH supervised the project, contributed to data interpretation, and provided critical revisions of the manuscript. NRP and SAS conceived the study, supervised the project, contributed to data interpretation, and provided critical revisions of the manuscript. NRP is the guarantor.

