## Supplement for "White Matter Integrity Correlates with Strength of Response to Deep Brain Stimulation in Treatment-Resistant Obsessive-Compulsive Disorder"

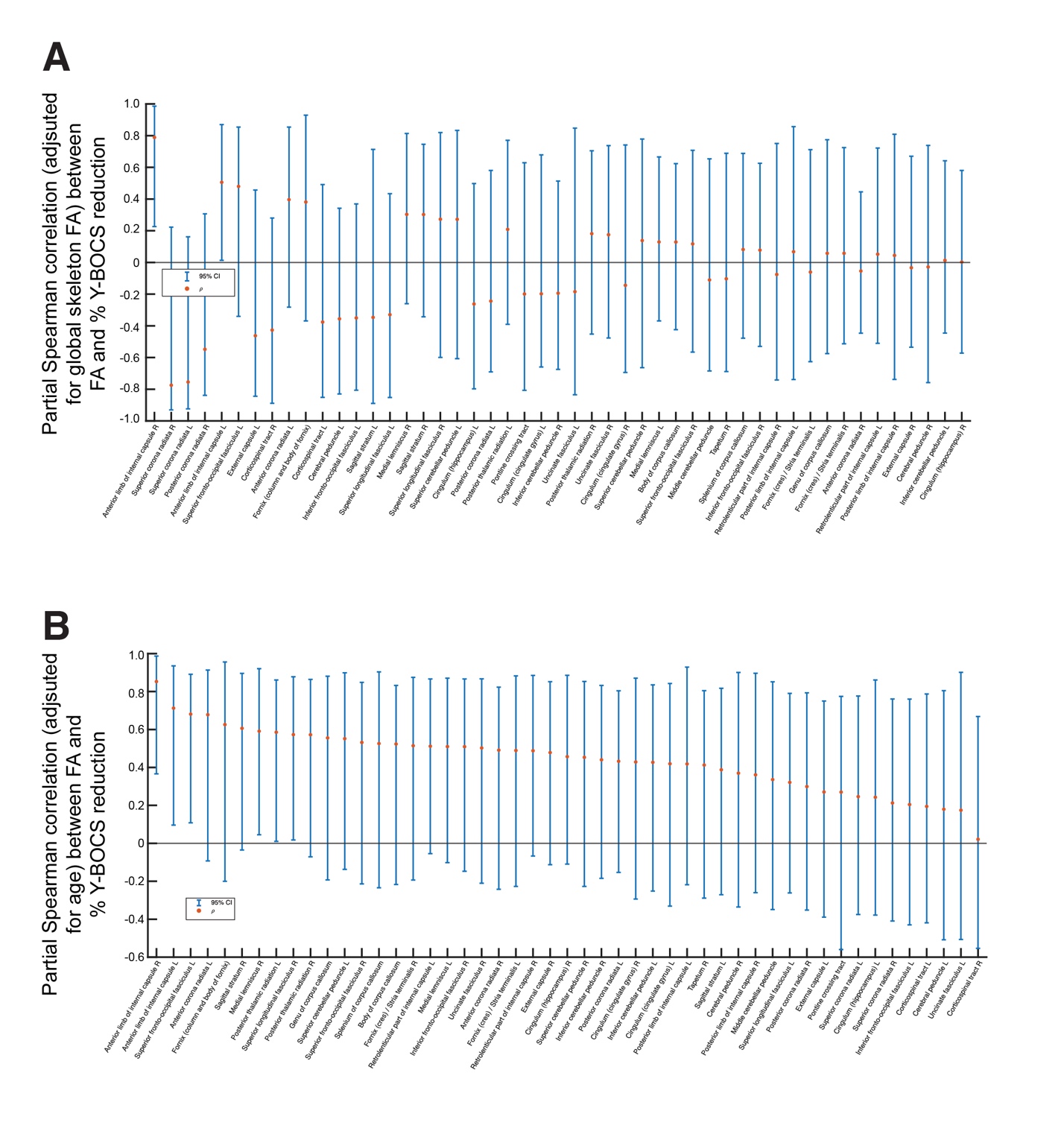

**Supplementary Figure 1: Partial Spearman correlation values for the relationship between % Y-BOCS reduction and FA controlling for global FA (A) and age (B).** The right ALIC has the highest partial correlation coefficient in both cases.
